# Quantifying selection bias due to unobserved patients in pharmacoepidemiologic studies of severe COVID-19 cohorts

**DOI:** 10.1101/2025.07.24.25332123

**Authors:** Marleen Bokern, Christopher T. Rentsch, Jennifer Quint, Anna Schultze, Ian Douglas

## Abstract

**Background:** The COVID-19 pandemic caused hospital pressures resulting in some patients with severe COVID-19 not being admitted. Studies aiming to measure treatment effects in patients with severe COVID-19 might produce biased estimates if restricted to hospitalised cohorts.

**Aim:** To quantify the effects of potential selection bias due to deaths outside of hospital in a case study of inhaled corticosteroids (ICS) and COVID-19 death among people with chronic obstructive pulmonary disease (COPD) hospitalised with COVID-19.

**Methods:** Using Clinical Practice Research Datalink Aurum linked to hospitalisation and death registries, we defined a cohort with COPD on 01 Mar 2020, followed up until 31^st^ August 2020. We assessed the odds of COVID-19 death (International Classification of Diseases, 10^th^ Revision U07) among hospitalised COVID-19 patients, comparing current users of ICS/long-acting β-agonist (LABA) and LABA/long-acting muscarinic antagonist (LAMA)). Our target population was those with COPD and severe COVID-19 warranting hospitalisation. We evaluated potential selection bias using quantitative bias analysis (QBA) in four plausible scenarios, varying assumed death rates among non-hospitalised patients. Selection probabilities for deaths due to COVID-19 were known. The assumptions were: (1) equal odds of death between non-hospitalised and hospitalised groups; (2) doubled odds of death in non-hospitalised ICS group compared to hospitalised; (3) halved odds of death in non-hospitalised ICS group; and (4) doubled odds of death in both treatment groups among non-hospitalised patients. We calculated bootstrapped 95% confidence intervals (CIs).

**Results:** During the study period, 107 ICS users and 133 LABA/LAMA users were hospitalised with COVID-19. COVID-19 deaths occurred in 42 (39.3%) ICS users versus 50 (37.6%) LABA/LAMA users. The OR after inverse probability of treatment weighting was 1.01 (95% CI 0.59-1.72). In scenario 1, the OR was unchanged (OR 1.07, 95% CI 0.70 – 1.67). In scenario 2, the corrected OR was 1.28 (95% CI 0.83 – 2.00). In scenario 3, the corrected OR was 0.81 (95% CI 0.52 – 1.23). In scenario 4, the corrected OR was 1.08 (95% CI 0.69 – 1.71).

**Conclusion:** QBA facilitated an assessment of the sensitivity of study results to potential selection bias. The results of the four scenarios presented are in line with the null hypothesis, but CIs were wide. Death rates in the non-hospitalised would have needed to be substantially different in the treatment groups to change the study conclusions.

## 1. Introduction

The COVID-19 pandemic led to an unprecedented surge in non-interventional studies. Many were designed to assess the impact of established and routinely prescribed medications for other indications to identify possible treatments for patients with severe COVID-19. These studies were often limited to hospitalised patients. However, treatment effects estimated within hospitalised cohorts can be affected by selection bias if conditioning is related to the exposure and outcome.^1^ Quantitative bias analysis (QBA) methods can provide a way to quantify the impact of biases. However, in practice, these methods are more rarely applied to account for selection bias than for confounding or misclassification.^2,3^

Inhaled corticosteroids (ICS) are commonly used maintenance medications in chronic obstructive pulmonary disease (COPD), which were investigated as potential repurposed drugs for the treatment of COVID-19 due to their immunosuppressive effects. These effects may have different consequences at different stages of COVID-19.^4^ Several observational studies investigated the impact of ICS on COVID-19 outcomes but found inconsistent results that may be affected by biases.^5–9^ RCTs consequently suggested a protective effect of initiating one ICS, budesonide, on severe COVID-19 outcomes in patients with mild COVID-19.^10,11^ Of note, observational studies often started with a population of patients hospitalised for COVID-19, indicating severe disease.^6,7^

Conditioning on an event such as hospitalisation for COVID-19 is a step we may take if we are interested in patients with severe COVID-19 as the target population, as hospitalisation can be viewed as a proxy for severe COVID-19. However, during the pandemic in the UK, there were hospital pressures and triaging processes that meant that some people with severe COVID-19 were not admitted to hospital.^12,13^ COVID-19 deaths occurred both in and outside hospitals, indicating that not everyone with severe COVID-19 was hospitalised.^14^ In this situation, investigators would be missing data on patients with severe COVID-19 who were never hospitalised. The extent of this will have varied over time depending on hospital pressures and COVID-19 prevalence. This may have led to selection bias in studies estimating treatment effects among hospitalised patients as some patients with severe COVID-19 were excluded. In this paper, we aim to quantify the effects of potential selection bias due to non-admission of severe COVID-19 cases in a study of ICS and COVID-19 death among people hospitalised for COVID-19.

## 2. Methods

The original study protocol was registered on ENCEPP EU PAS (register number 47885).

## Data

The data sources, study population, and exposure, outcome and covariate definitions have previously been described^15^, and are summarised below.

### Data source

This study used routinely primary care collected data from the UK recorded in Clinical Practice Research Datalink (CPRD) Aurum. CPRD Aurum includes data on 41 million patients (May 2022 build), with over 13 million patients currently registered (20% of the UK population)^16^ from >1,300 general practices (GPs) which use EMIS GP patient management software.^11^ CPRD Aurum is broadly representative of the English population.^17^

CPRD Aurum was linked to Hospital Episode Statistics (HES) Admitted Patient Care (APC) and Office for National Statistics (ONS) Death Registry.^17,18^ HES APC holds information on all in-patient contacts at NHS hospitals in England.^18,19^ The ONS death registry contains information on deaths in England and Wales, including a cause of death documented using International Classification of Disease 10^th^ revision (ICD-10) codes.^19,20^ Data was also linked to the Index of Multiple Deprivation (IMD), a postcode-level indicator of socioeconomic status.

### Study population

We defined a cohort of people diagnosed with COPD before 01^st^ March 2020 (i.e., baseline) based on a validated algorithm to identify COPD in CPRD.^21^ Patients were required to be alive and registered in CPRD Aurum at baseline, with at least 12 months’ continuous registration prior to baseline. In accordance with National Institute for Health and Care Excellence (NICE) guidelines for COPD diagnosis^33^, patients needed to be aged ≥35 and have a record of current or former smoking at any point before baseline. Patients were included if they had a COVID-19 hospitalisation, determined using ICD-10 U07.1/U07.2 as a primary diagnosis code in HES APC. We excluded people with asthma diagnoses recorded within three years before baseline, leukotriene receptor antagonist use within 4 months before baseline, as this indicates asthma, or other chronic respiratory disease at any point before baseline. The admission date for first COVID-19 hospitalisation was the start of follow-up (index date). Patients were followed up until death (recorded in ONS or CPRD), deregistration, or 31^st^ August 2020 (end of first pandemic wave), whichever came first. If death was registered in ONS, we used that date as the date of death. If death was missing in ONS but registered in CPRD, we used the date recorded in CPRD as the date of death. A study diagram^22^ is in Supplementary Figure 1.

#### Exposure

Continuous treatment episodes were generated based on the recorded prescription issue date and information on the intended duration, prescribed amount and dosage.^15^

We used the derived treatment episodes to categorise people as exposed to ICS/LABA or LABA/LAMA on 01 March 2020 as combined or separate inhalers. People using ICS/LABA/LAMA (i.e., triple therapy) were excluded as we expected patients using triple therapy to be sicker than those on dual therapy. ICS/LABA was the exposure of interest and LABA/LAMA was the active comparator.

#### Outcome

The outcome was death with COVID-19 (U07.1 and U07.2) as a cause of death anywhere on the death certificate in the ONS Death Registry.

### Analysis

#### Baseline characteristics and logistic regression models

Cohort characteristics were summarised using descriptive statistics by exposure group. The following covariates, selected based on input from a practising clinician, were assessed: age, sex, body mass index (BMI, most recent within 10 years, categorised as underweight (<18.5), normal (18.5-24.9), overweight (25-29.9), or obese (≥30)), smoking (current vs former), ethnicity, cancer (ever), diabetes (ever), chronic kidney disease (ever), cardiovascular disease (ever), hypertension (ever), asthma (>3 years prior to baseline), immunosuppression, influenza vaccination (past year), pneumococcal vaccination (past 5 years), IMD quintile, and COPD exacerbations in the past 12 months, based on a validated algorithm.^25^

There were missing data for BMI, ethnicity and IMD. Missing BMI was assumed to be normal in line with previous work.^23^ Missing ethnicity was treated as a separate category^24^, and patients with missing IMD were excluded as this may indicate poor quality records.

We estimated propensity scores (PSs) and used inverse probability of treatment weighting (IPTW) to estimate the average treatment effect (ATE) adjusting for potential confounders among those hospitalised for COVID-19. PSs were estimated using logistic regression including the covariates listed above. Weights were calculated as 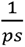 (ICS/LABA) and 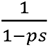 (LABA/LAMA), where the PS is the probability of receiving ICS. To ensure that the inverse probability of treatment (IPT)-weighted pseudopopulation had the same total sample size as the observed population, we applied k-scaling: weights were multiplied by a constant factor k, defined as the ratio of the observed sample size to the total weighted sample size. This scaling forces the weighted sample size to equal the original sample size and avoids reporting non-integer patient counts. Overlap of the PSs across treatment groups was assessed graphically and by summarising PSs by treatment group. PSs were trimmed to the region of common support.^26^

Logistic regression models were used to estimate odds ratios (ORs) and 95% confidence intervals (CIs), both unweighted and IPT-weighted.

#### Selection bias

Selection into the study followed the flowchart in Figure 1, which illustrates the situation in a typical electronic health record (EHR) study using population-level data from primary care, hospitalisations and death registries. The target population was people with COVID-19 severe enough to warrant hospitalisation. When defining the study population as people who were hospitalised, patients in the dark blue boxes could be observed while the light blue boxes were unknown. We observed the number of people in the general population exposed to ICS or LABA/LAMA, those who became hospitalised, and those who died with or without hospitalisation. If conditioning on hospitalisation leads to systematic differences in terms of outcome risk in the type of patients selected from the ICS group compared to those selected from the LABA/LAMA group, selection bias may arise due to differences in outcome risk. Differences could arise for various reasons, for example if the occurrence of severe COVID-19 and hospitalisation depends on unobserved factors such as frailty and care home residence^27^ that are unequally distributed between treatment groups. A directed acyclic graph (DAG) depicting the assumed structure of selection bias is in Supplementary Figure 2.

**Figure 1.**
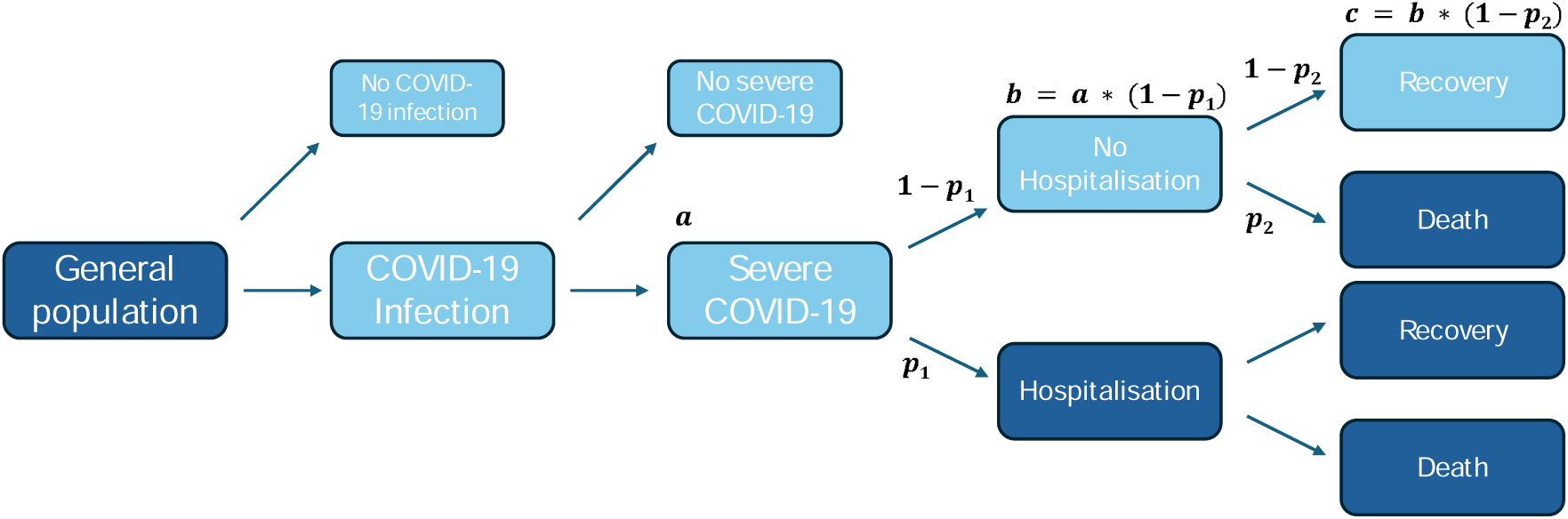
Typical selection pathway in an electronic health record study investigating COVID-19. Dark blue boxes can be observed from data, while light blue boxes remain unobserved. a = number of patients with severe COVID-19, p_1_ = probability of hospitalisation among those with severe COVID-19, b = number of patients with severe COVID-19 who were never hospitalised, p_2_ = probability of death among those with severe COVID-19 who were not hospitalised, c = number of patients with severe COVID-19 who recovered without hospitalisation.

We assumed that all people who were hospitalised or died with COVID-19 had severe COVID-19 and that those with severe COVID-19 who were not hospitalised had similar COVID-19 severity to those hospitalised. Given this, we used the observed odds of death among the hospitalised to set a range of plausible parameters for the odds of death among the non-hospitalised in each treatment group.

#### Correcting for selection bias

Classical correction for selection bias uses the selection probabilities for each combination of exposure and outcome among the target population, which are used to calculate a selection bias odds ratio (sOR) which is applied to the observed effect estimate.^28,29^

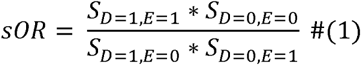

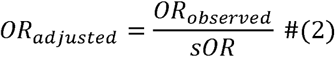

S is the selection probability, D is the outcome, and E the exposure. In this case, the target population was unobserved, so not all selection probabilities could be directly estimated from the data. In these situations, plausible estimates for selection probabilities in each treatment group could be derived from published literature, clinical experience or representing best- or worst-case scenarios to evaluate the sensitivity of the findings to selection pressures.

In our scenario, plausible ranges for the selection probabilities could be calculated after estimating the hospitalisation probability among those with severe COVID-19 (p_1_, Figure 1), the number of people with severe COVID-19 (a), the number of people with severe COVID-19 not hospitalised (b), or the probability or odds of death outside of hospital (p_2_). As we had data on deaths without preceding hospitalisation, we could calculate the selection probabilities for COVID-19 deaths and needed to estimate only the selection probabilities for the recovered.

As we observed the odds of death among the hospitalised, we estimated the odds of death among the non-hospitalised by treatment group. This allowed us to calculate the number of recovered non-hospitalised patients as

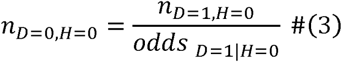

N represents numbers of patients, D is the outcome, and H is hospitalisation. We then calculate the number of people with severe COVID-19 by treatment group by adding the unobserved number of recovered non-hospitalised patients *n_D_*_=0,*H*=0_ to the observed total deaths and the observed number of hospitalised recovered. This allows us to calculate the selection probabilities and to correct the observed OR for selection bias (Equation 1).

We calculated the ORs varying the odds of death among the non-hospitalised between 0.05 and 2.0 in each treatment group in 0.05 increments. Odds of death among the hospitalised are known from the data. We depict the results of this analysis with a heat map.

Additionally, we highlight 4 scenarios assuming odds of death among the non-hospitalised. 95% CIs were estimated using bootstrapping with 10,000 iterations.

##### Scenario 1: Non-hospitalised same as hospitalised

The odds of death, by treatment group, are the same among those who were not hospitalised as among those who were hospitalised. This means that selection probabilities were assumed to be equal for unobserved exposed and unexposed non-cases. This represents a situation where hospitalisation had no impact on survival and theoretically, we would expect this to result in zero bias.

##### Scenario 2: ICS non-hospitalised sicker than LABA/LAMA non-hospitalised

The odds of death in the LABA/LAMA group are the same among those who were not hospitalised as among those who were hospitalised but are doubled in the ICS group among those not hospitalised compared with those hospitalised. This models a scenario where non-hospitalised ICS patients are sicker than non-hospitalised LABA/LAMA patients. Alternatively, it can be thought of as setting higher selection probabilities for exposed non-cases compared to unexposed non-cases.

##### Scenario 3: ICS non-hospitalised healthier than LABA/LAMA non-hospitalised

The odds of death in the LABA/LAMA group are the same among those who were not hospitalised as among those who were hospitalised, but are halved in the ICS group among those not hospitalised compared with those hospitalised. This models a scenario where non-hospitalised ICS patients are healthier than non-hospitalised LABA/LAMA patients.

##### Scenario 4: Non-hospitalised sicker than hospitalised

The odds of death are doubled among those who were not hospitalised compared to those who were hospitalised regardless of treatment group.

As a diagnostic check, we calculated the other unknown quantities in Figure *1* for scenarios 1-4 by treatment group. The total number of non-hospitalised patients with severe COVID-19 was calculated by summing the previously calculated number of patients with severe COVID-19 who were not hospitalised and recovered and the total number of COVID-19 deaths outside of hospital.

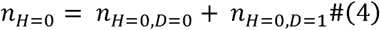

Thereupon, the total number of people with severe COVID-19 was calculated as the calculated number of non-hospitalised patients with severe COVID-19 plus the hospitalised.

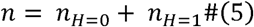

Finally, the corresponding probability of hospitalisation was calculated as the hospitalised divided by the assumed total number of patients with severe COVID-19.

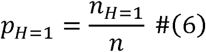

Data was managed using Stata MP version 17.0^30^ and analysis carried out using R (version 4.4.2)^31^. Code lists and data management and analysis code are on GitHub (https://github.com/bokern/ics_covid_collider).

## 3. Results

### Analysis restricting to hospitalised patients

The COPD cohort consisted of 14,906 (40.0%) ICS users and 22,319 (60.0%) LABA/LAMA users. Of those, 107 ICS users and 133 LABA/LAMA users were hospitalised with COVID-19 and were included in this analysis (Table *1*). In the ICS group, 42 (39.3%) experienced the outcome COVID-19 death, compared with 50 (37.6%) in the LABA/LAMA group.

**Table 1.**
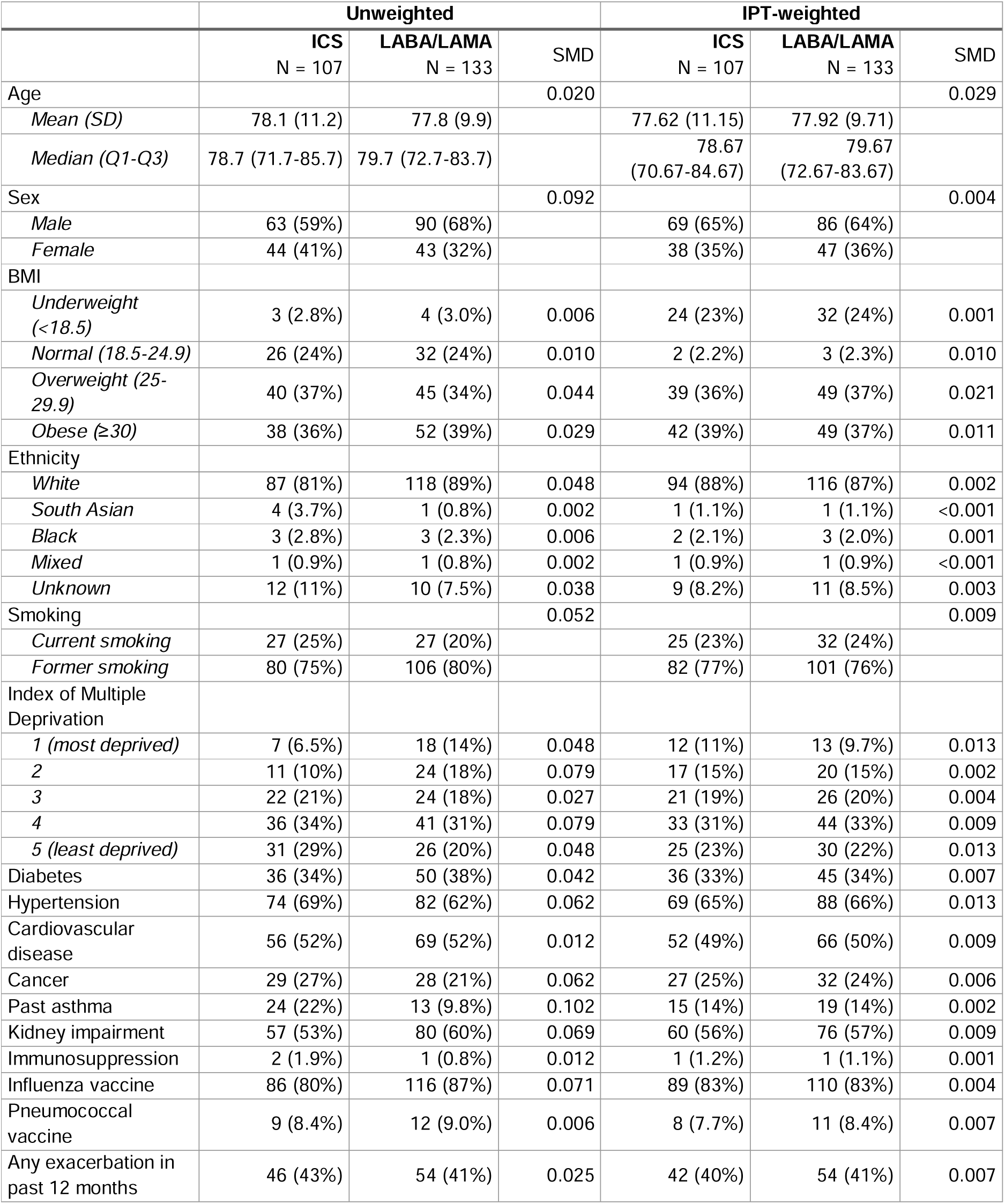
baseline cohort characteristics before and after inverse probability of treatment weighting. BMI = Body mass index, IPT = inverse probability of treatment, SD = standard deviation, SMD = standardised mean difference

|  | Unweighted |  |  | IPT-weighted |  |  |
| --- | --- | --- | --- | --- | --- | --- |
|  | ICS<br>N = 107 | LABA/LAMA<br>N = 133 | SMD | ICS<br>N = 107 | LABA/LAMA<br>N = 133 | SMD |
| Age |  |  | 0.020 |  |  | 0.029 |
| *Mean (SD)* | 78.1 (11.2) | 77.8 (9.9) |  | 77.62 (11.15) | 77.92 (9.71) |  |
| *Median (Q1-Q3)* | 78.7 (71.7-85.7) | 79.7 (72.7-83.7) |  | 78.67<br>(70.67-84.67) | 79.67<br>(72.67-83.67) |  |
| Sex |  |  | 0.092 |  |  | 0.004 |
| *Male* | 63 (59%) | 90 (68%) |  | 69 (65%) | 86 (64%) |  |
| *Female* | 44 (41%) | 43 (32%) |  | 38 (35%) | 47 (36%) |  |
| BMI |  |  |  |  |  |  |
| *Underweight (<18.5)* | 3 (2.8%) | 4 (3.0%) | 0.006 | 24 (23%) | 32 (24%) | 0.001 |
| *Normal (18.5-24.9)* | 26 (24%) | 32 (24%) | 0.010 | 2 (2.2%) | 3 (2.3%) | 0.010 |
| *Overweight (25-29.9)* | 40 (37%) | 45 (34%) | 0.044 | 39 (36%) | 49 (37%) | 0.021 |
| *Obese (≥30)* | 38 (36%) | 52 (39%) | 0.029 | 42 (39%) | 49 (37%) | 0.011 |
| Ethnicity |  |  |  |  |  |  |
| *White* | 87 (81%) | 118 (89%) | 0.048 | 94 (88%) | 116 (87%) | 0.002 |
| *South Asian* | 4 (3.7%) | 1 (0.8%) | 0.002 | 1 (1.1%) | 1 (1.1%) | <0.001 |
| *Black* | 3 (2.8%) | 3 (2.3%) | 0.006 | 2 (2.1%) | 3 (2.0%) | 0.001 |
| *Mixed* | 1 (0.9%) | 1 (0.8%) | 0.002 | 1 (0.9%) | 1 (0.9%) | <0.001 |
| *Unknown* | 12 (11%) | 10 (7.5%) | 0.038 | 9 (8.2%) | 11 (8.5%) | 0.003 |
| Smoking |  |  | 0.052 |  |  | 0.009 |
| *Current smoking* | 27 (25%) | 27 (20%) |  | 25 (23%) | 32 (24%) |  |
| *Former smoking* | 80 (75%) | 106 (80%) |  | 82 (77%) | 101 (76%) |  |
| Index of Multiple Deprivation |  |  |  |  |  |  |
| *1 (most deprived)* | 7 (6.5%) | 18 (14%) | 0.048 | 12 (11%) | 13 (9.7%) | 0.013 |
| *2* | 11 (10%) | 24 (18%) | 0.079 | 17 (15%) | 20 (15%) | 0.002 |
| *3* | 22 (21%) | 24 (18%) | 0.027 | 21 (19%) | 26 (20%) | 0.004 |
| *4* | 36 (34%) | 41 (31%) | 0.079 | 33 (31%) | 44 (33%) | 0.009 |
| *5 (least deprived)* | 31 (29%) | 26 (20%) | 0.048 | 25 (23%) | 30 (22%) | 0.013 |
| Diabetes | 36 (34%) | 50 (38%) | 0.042 | 36 (33%) | 45 (34%) | 0.007 |
| Hypertension | 74 (69%) | 82 (62%) | 0.062 | 69 (65%) | 88 (66%) | 0.013 |
| Cardiovascular disease | 56 (52%) | 69 (52%) | 0.012 | 52 (49%) | 66 (50%) | 0.009 |
| Cancer | 29 (27%) | 28 (21%) | 0.062 | 27 (25%) | 32 (24%) | 0.006 |
| Past asthma | 24 (22%) | 13 (9.8%) | 0.102 | 15 (14%) | 19 (14%) | 0.002 |
| Kidney impairment | 57 (53%) | 80 (60%) | 0.069 | 60 (56%) | 76 (57%) | 0.009 |
| Immunosuppression | 2 (1.9%) | 1 (0.8%) | 0.012 | 1 (1.2%) | 1 (1.1%) | 0.001 |
| Influenza vaccine | 86 (80%) | 116 (87%) | 0.071 | 89 (83%) | 110 (83%) | 0.004 |
| Pneumococcal vaccine | 9 (8.4%) | 12 (9.0%) | 0.006 | 8 (7.7%) | 11 (8.4%) | 0.007 |
| Any exacerbation in past 12 months | 46 (43%) | 54 (41%) | 0.025 | 42 (40%) | 54 (41%) | 0.007 |

Using logistic regression, the unweighted OR was 1.07 (95% CI 0.63–1.81), which moved towards the null after IPTW (OR 1.01 (95% CI 0.59-1.72)).

**Figure 2.**
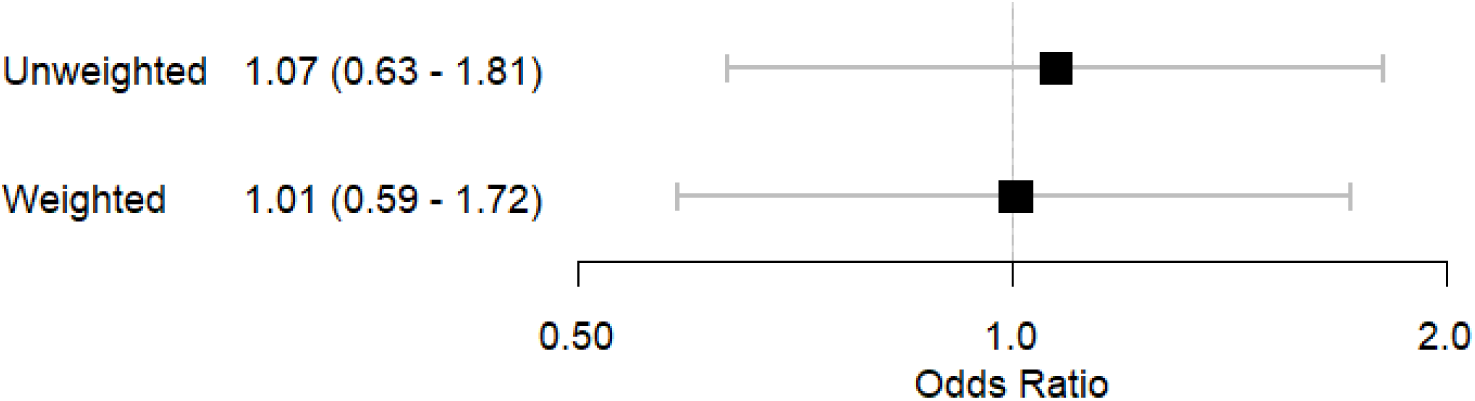
Results of logistic regression models with 95% confidence intervals before and after inverse probability of treatment weighting

### Selection bias

There were 20 deaths without hospitalisation in the ICS group and 22 in the LABA/LAMA group (Figure *3*). Among those hospitalised, the odds of death were 0.65 in the ICS group and 0.60 in the LABA/LAMA group.

**Figure 3.**
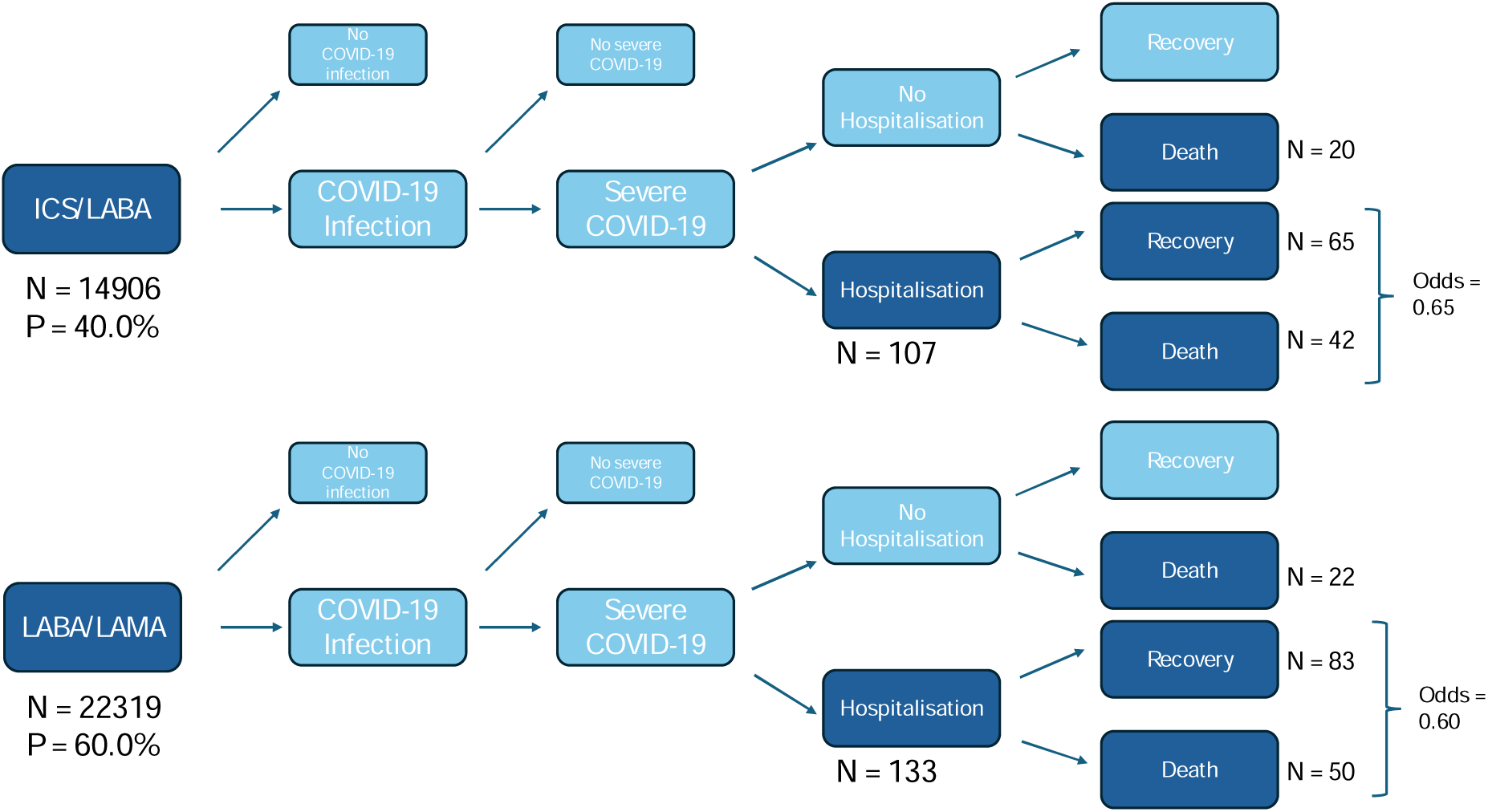
Flowchart of patient selection. “Severe COVID-19” means COVID-19 severe enough to warrant hospitalisation. Dark blue boxes represent populations we can observe from the available data. Light blue boxes cannot be observed.

**Error! Reference source not found.** presents the results of scenarios 1-4. ORs vary between 0.81 and 1.27, with all 95% CIs crossing the null (Figure 4), with corresponding probabilities of hospitalisation among those with severe COVID-19 between 0.57 – 0.77 (Supplementary Table 1). When varying the assumed odds of death incrementally, the ORs for most combinations of odds of death were similar to the observed OR, meaning the majority (54%) of ORs were within 0.2 of the observed OR (Figure 5).

**Figure 4.**
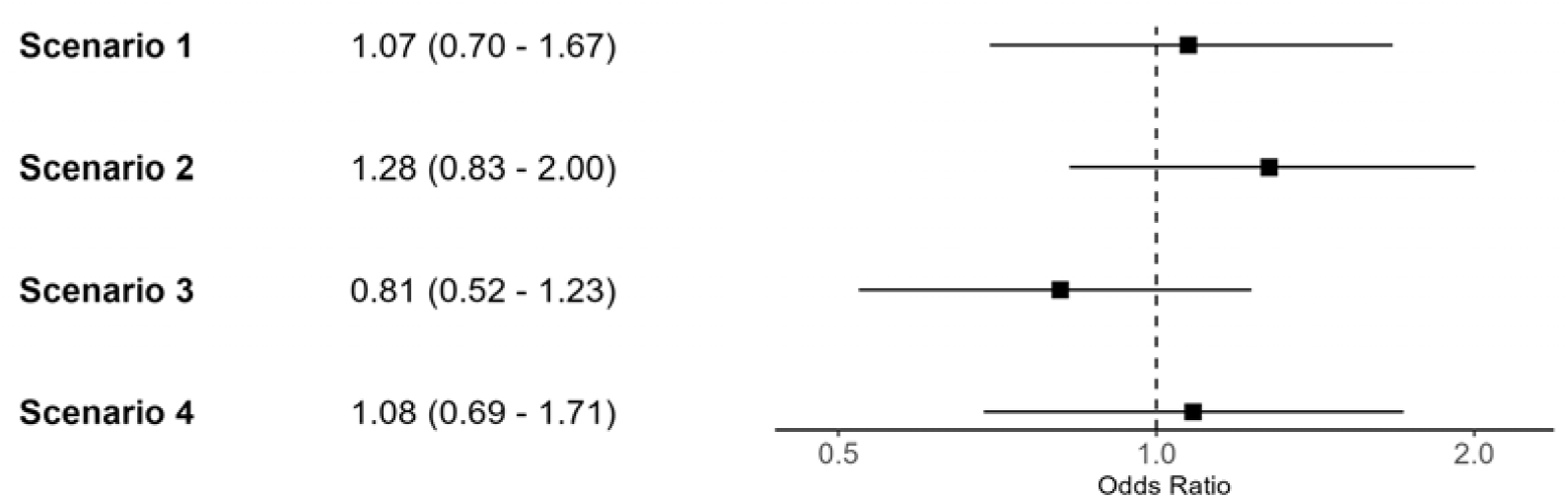
Forest plot of odds ratios and 95% confidence intervals after correction for selection bias. CIs were generated using bootstrapping with 10000 iterations.

**Figure 5.**
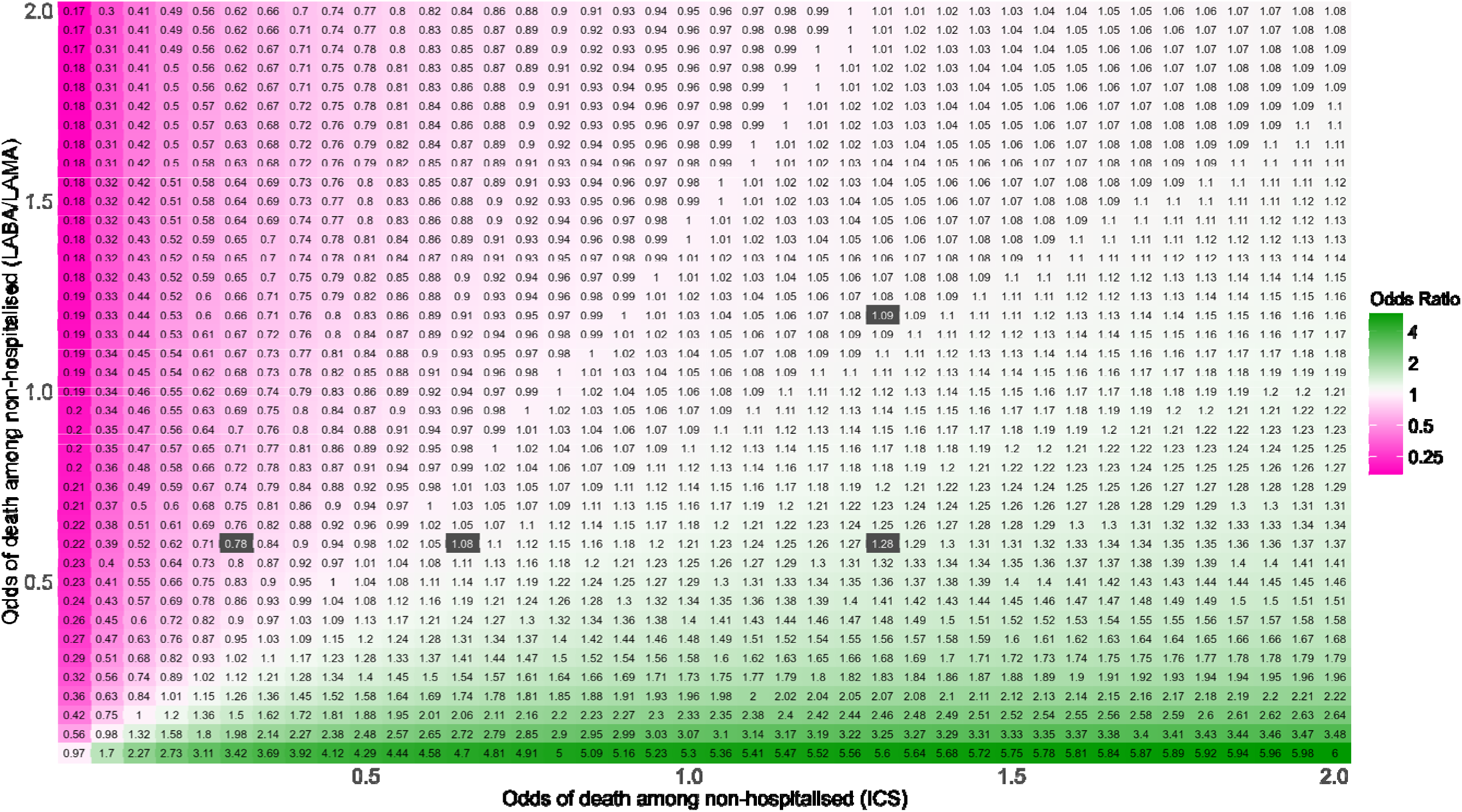
Heat map depicting odds ratios corrected for differential odds of death among the (unobserved) non-hospitalised patients with severe COVID-19. The colour scheme is centred around the odds ratio (OR) observed when restricting to hospitalised patients (OR = 1.08). Pink represents a decrease in OR compared to the observed, green represents an increase in OR compared to the observed. The four dark grey boxes represent scenarios 1-4. Scenario 1: odds of death among the non-hospitalised are the same as among the hospitalised (OR = 1.073, bottom middle), scenario 2: odds in LABA/LAMA group are constant, odds in the ICS group are doubled among the unobserved (OR = 1.279, bottom right), scenario 3: odds in LABA/LAMA group are constant, odds in the ICS group are halved among the unobserved (OR = 0.811, bottom left), scenario 4: odds of death are doubled in both groups (OR = 1.0835, top right). Any differences between depicted ORs and calculated ORs are due to rounding.

**Table 2.**
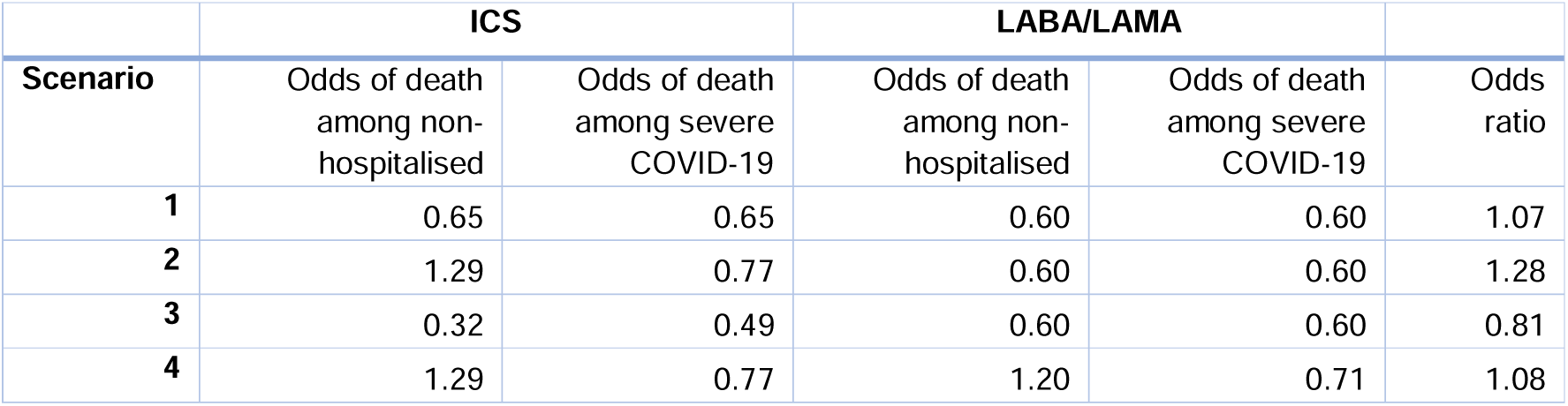
Results of scenarios 1-4.

## 4. Discussion

We have applied a method to account for potential selection bias in a study of COVID-19 mortality estimating treatment effects among individuals with severe COVID-19 using a hospitalised cohort. After correcting for the potential selection bias, the adjusted OR for COVID-19 death comparing ICS users with LABA/LAMA users was similar to the observed OR for many combinations of selection probabilities. For four pre-defined plausible scenarios, ORs varied between 0.81 when the odds of death among the unobserved in the ICS group were halved compared with the observed, and 1.28 when the odds of death among the unobserved in the ICS group were doubled compared with the observed. However, 95% CIs calculated using bootstrapping overlapped with a null effect under all scenarios. For the conclusions to have significantly differed from those of the observed results, the odds of death among unobserved ICS patients would have needed to be either less than half or more than double compared to the observed.

### Comparison with Previous Work

Selection bias has not frequently been addressed in pharmacoepidemiologic studies.^2,3^ Some examples come from the field of perinatal pharmacoepidemiology. A study investigating selective serotonin reuptake inhibitor use in pregnancy and cardiac defects assessed potential selection bias due to missing terminations, correcting the observed OR using selection probabilities for each exposure-outcome combination.^32^ Further studies investigating lithium use and cardiac malformations and statins and congenital malformations used similar methods.^29,33^ One study investigated selection bias in a study of allergy medications and COVID-19 testing.^34^ The authors used a bounding factor^35^ to calculate the smallest the true OR could be given assumed risk (or odds) ratios between a selection factor and the exposure and outcome. This method can be used when correcting for collider bias where the selected population is the target population.^34,35^

Regarding the clinical question, two previous studies investigating the effects of ICS on COVID-19 outcomes among hospitalised patients, both suggesting no effect of ICS. In a cohort of COPD patients hospitalised with COVID-19 in Denmark, routine use of ICS was not associated with an increased risk of death (HR 1.02 (95% CI, 0.78–1.32)) compared to bronchodilator use. In a hospitalised UK cohort, among those with chronic pulmonary disease, both those with and without routine ICS use had an increased hazard of death with COVID-19 (no ICS use: HR 1.16, 95% CI 1.12-1.22; using ICS: HR 1.10, 95% CI 1.04-1.16) compared to patients without respiratory disease.^7^ As this study was also set in the UK during the first wave of the pandemic, selection pressures among people with respiratory diseases would have been similar to this study. However, because the comparator group did not have respiratory disease, the findings are not directly comparable to ours.

### Interpretation

We illustrate that selection bias is introduced if there are differential selection pressures on the exposed and unexposed groups. This may be less likely in pharmacoepidemiological studies using an active comparator and is confirmed with very similar selection probabilities for cases in the exposed and unexposed groups (0.68 in the ICS group and 0.69 in the LABA/LAMA group).

While the point estimates vary significantly when varying selection probabilities, differences in selection probabilities for the non-cases as large as assumed in scenarios 2 and 3 may be unrealistic given the treatment groups were relatively similar (Table *1*). Furthermore, one may assume that hospitalisation would reduce the risk of death compared to those with severe COVID-19 who were not admitted, making scenarios 2 and 4 potentially more likely than scenarios 1 and 3. For all four calculated scenarios, the CIs are in line with the null hypothesis and the uncorrected result.

For a mechanism to introduce bias, either the selection probabilities for non-cases need to differ for exposed and unexposed individuals or, equivalently, the recovery rates for the non-hospitalised need to differ between treatment groups. Hospital pressures during the pandemic meant that some people with severe COVID-19 may not have been admitted to hospital if they were deemed unlikely to survive, as resources would have been preferentially allocated to patients with higher survival probability.^36^

We used an active comparator design to minimise differences between our comparison groups. Traditionally, this approach is used to deal with confounding by indication. A further benefit of use of active comparators is to reduce differences in selection probabilities by ensuring comparison groups have similar background illnesses and are therefore treated more similarly in a health and social care system.

They can therefore also help mitigate the risk of selection bias.^37^ Much of the literature on QBA for selection bias does not take active comparators into account.^1,38^

In this study, the probability of being hospitalised with COVID-19 may depend on place of residence (e.g., care home vs private homes) and frailty. In these datasets, care home residence is not readily available, and frailty can be difficult to ascertain in EHRs.^39^ Hospital admissions from care homes dropped at the start of the pandemic.^40^ Additionally, people with COVID-19 were discharged to care homes, causing outbreaks within care homes.^41^ Taken together, people living in care homes may have been less likely to be admitted to hospital with severe COVID-19 and therefore may be more likely to be missed when restricting to hospitalisation. Such a mechanism could underpin different probabilities of hospital admission between comparison groups, and the extent of these differences would then influence the degree to which selection bias may lead to incorrect results.

### Strengths and Limitations

Strengths of this study include the representativeness of CPRD Aurum, as well as the comprehensive capture of hospitalisations and deaths.

QBA is not frequently used in non-interventional research, and QBA for selection bias is particularly rare.^2,3^ This may be because quantifying selection probabilities is difficult in many situations. We have illustrated how these methods can be implemented with reference to odds of the outcome amongst the exposed, as opposed to sampling probabilities, which may be easier for researchers to estimate. Illustrating the selection process using a flowchart requires researchers to think through selection processes and may further aid in the application of QBA. In the absence of well-informed estimates of selection probabilities, heat maps can show a wide range of assumptions simultaneously.

The correction method presented here does not attempt to account for residual confounding. Doing so would require more complex modelling or simulation methods and therefore not be pragmatic to implement for many researchers. Alternatively, selection probabilities or odds of death can be estimated within strata of confounders.^42^ However, conventional analyses weighted by PS showed little change in OR both when restricting to hospitalised patients, and in the whole COPD cohort.^15^ While this is not proof that confounding did not affect analyses among people with severe COVID-19, this is reassuring regarding the suitability of the active comparator.

The flowchart (**Error! Reference source not found.**) is specific to this time period in the UK when COVID-19 testing was limited, and we therefore treat COVID-19 infections as missing. In other settings where virus circulation is lower or testing more widely available, test data may provide a reliable estimate of infections, and therefore of those susceptible to severe COVID-19. In an acute pandemic scenario, it may also be more feasible to assess the selection probabilities by making assumptions on probability of infection and severe disease rather than survival outcomes. The choice of parameters to estimate may depend on the nature of the selection process and what data is available at different time points.

Our approach of calculating recovery rates relies on data on deaths outside of hospitals, so selection probabilities for people with the outcome were known. This may be the case if the outcome is ascertained through linked data. If these probabilities were unknown, more assumptions would have to be made (e.g. odds of death among the non-hospitalised, and the number of either deaths or recoveries outside of hospital by treatment group).

We focus on selection bias due to missing data. However, even if all severe COVID-19 cases were hospitalised and selected into the study, collider bias may still distort associations between factors that cause hospitalisation and should be dealt with as a separate consideration.^1,27^ Methods to account for selection bias in this special case where the target population is the selected population have overlap with methods to adjust for confounding.^43^ We chose an active comparator that minimises confounding, which simultaneously reduces selection bias. Additionally, as is the case for confounding, methods exist to bound selection bias.^35^ Based on Equation 6, the upper bound for the selection probabilities are equal to the number of hospitalisations divided by the number of hospitalisations plus the number of deaths outside of hospital. For the ICS group, the upper bound for the probability of hospitalisation among those with severe COVID-19 was 0.843, and for the LABA/LAMA group, it was 0.858.

In summary, we demonstrate the use of QBA to correct for assumed differential selection probabilities in a pharmacoepidemiological study. While all QBA is based on assumptions, our study highlights the value of attempting to quantify the potential impact of selection bias. In this example, we demonstrated that plausible differences in selection probabilities were unlikely to have changed the conclusions of the study.

## 5. Declarations

## Data Availability

No additional data is available. Data management and analysis code, along with all code lists, are available on our GitHub repository (https://github.com/bokern/ics_covid_collider).

## Acknowledgments

This study is based in part on data from the Clinical Practice Research Datalink obtained under licence from the UK Medicines and Healthcare products Regulatory Agency. The data is provided by patients and collected by the NHS as part of their care and support. The interpretation and conclusions contained in this study are those of the author/s alone. The study was approved by the Independent Scientific Advisory Committee (approval number: 22_001876).

## Competing Interests

MPB is funded by a GSK PhD studentship to investigate the application of quantitative bias analysis in observational studies of COVID-19. IJD has unrestricted grants from and shares in GSK. AS is employed by LSHTM on a fellowship funded by GSK. CTR reports no conflicts of interest.

## Funding

Marleen Bokern is funded by a GSK PhD studentship to undertake this work.

## Ethics Approval

The study was approved by the London School of Hygiene and Tropical Medicine Research Ethics Committee (Reference: 27896) and the Independent Scientific Advisory Committee of the UK Medicines and Healthcare Products Regulatory Agency (Approval Number: 22_001876).

## Consent for publication

Not applicable.

## Author contributions

MB, AS, CTR, IJD contributed to the study design. MB conducted the data management and analysis and drafted the manuscript. All authors contributed to reviewing and editing of the manuscript. All authors were involved in design and conceptual development and reviewed and approved the final manuscript.

## Notes

### Competing Interest Statement

MB is funded by a GSK PhD studentship to investigate the application of quantitative bias analysis in observational studies of COVID-19. ID has unrestricted grants from and shares in GSK. AS is employed by LSHTM on a fellowship funded by GSK. CTR and JQ report no conflicts of interest.  

