## Supplement for "Quantifying selection bias due to unobserved patients in pharmacoepidemiologic studies of severe COVID-19 cohorts"

**Supplementary Figure 1 Study diagram**

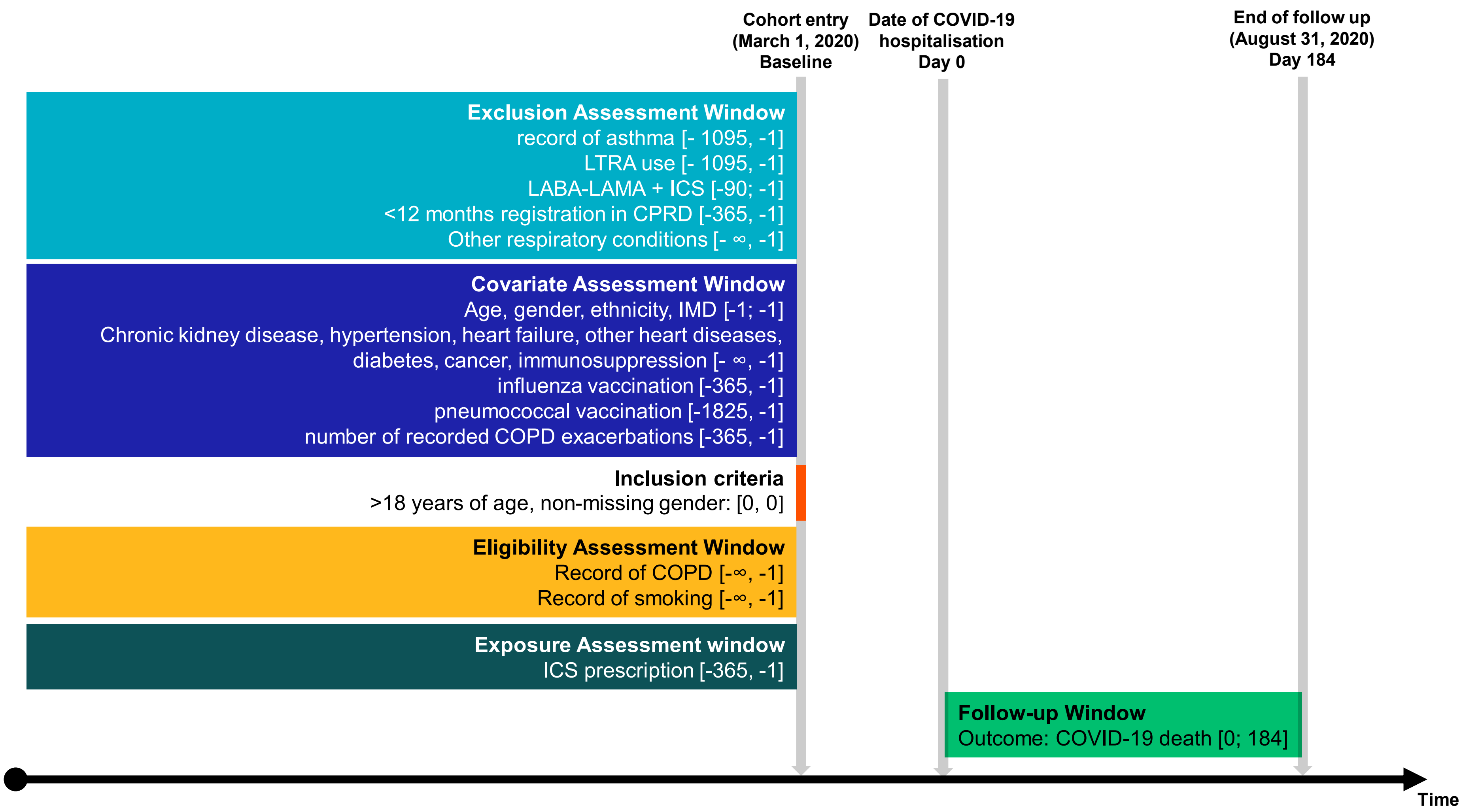

**Supplementary Figure 2 Directed acyclic graph (DAG) depicting the assumed structure of selection bias**

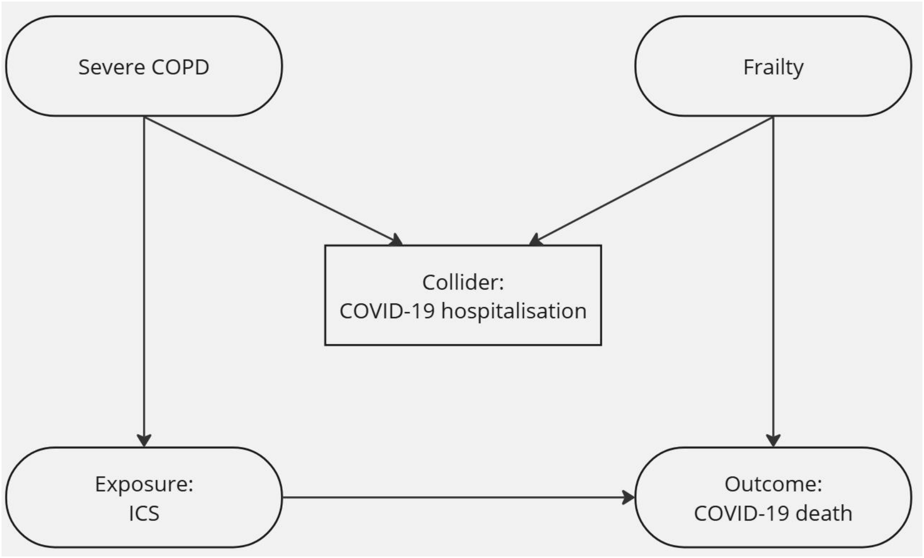

**Supplementary Figure 3 Unweighted propensity score distribution**

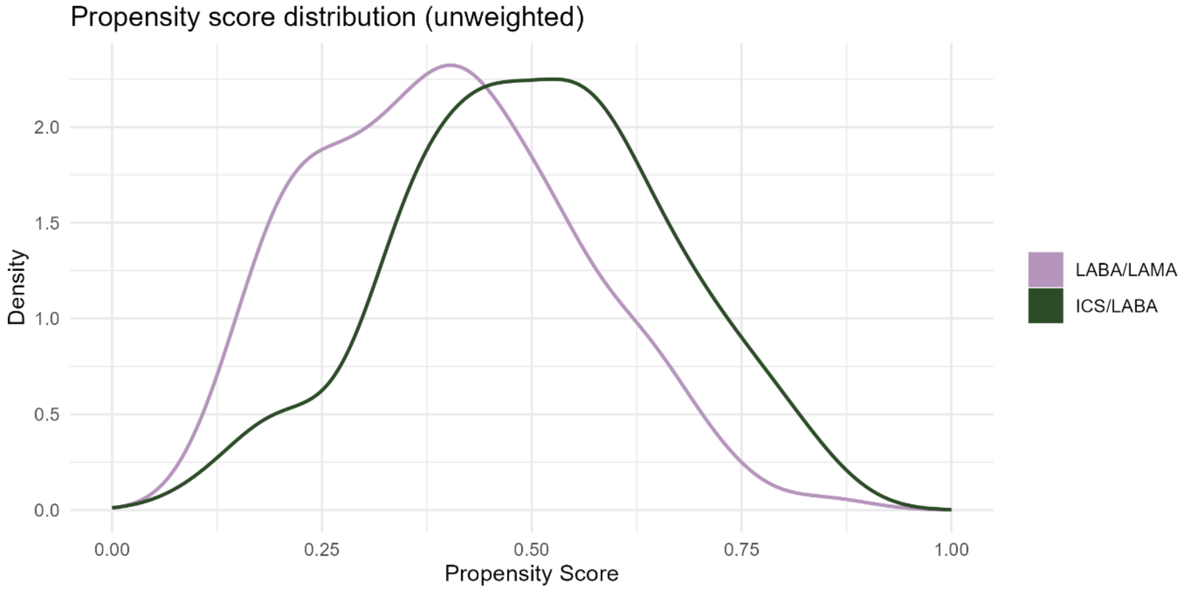

**Supplementary Figure 3 Propensity score distribution after inverse probability of treatment weighting**

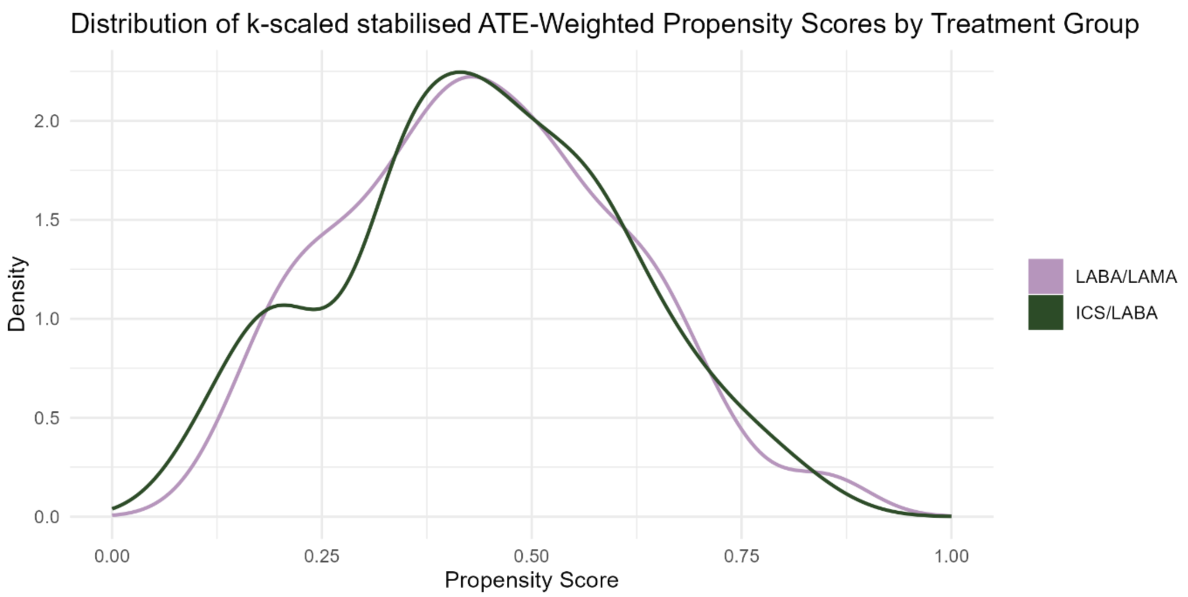

**Supplementary Figure 3 Absolute standardised mean differences (SMDs) before and after inverse probability of treatment weighting**

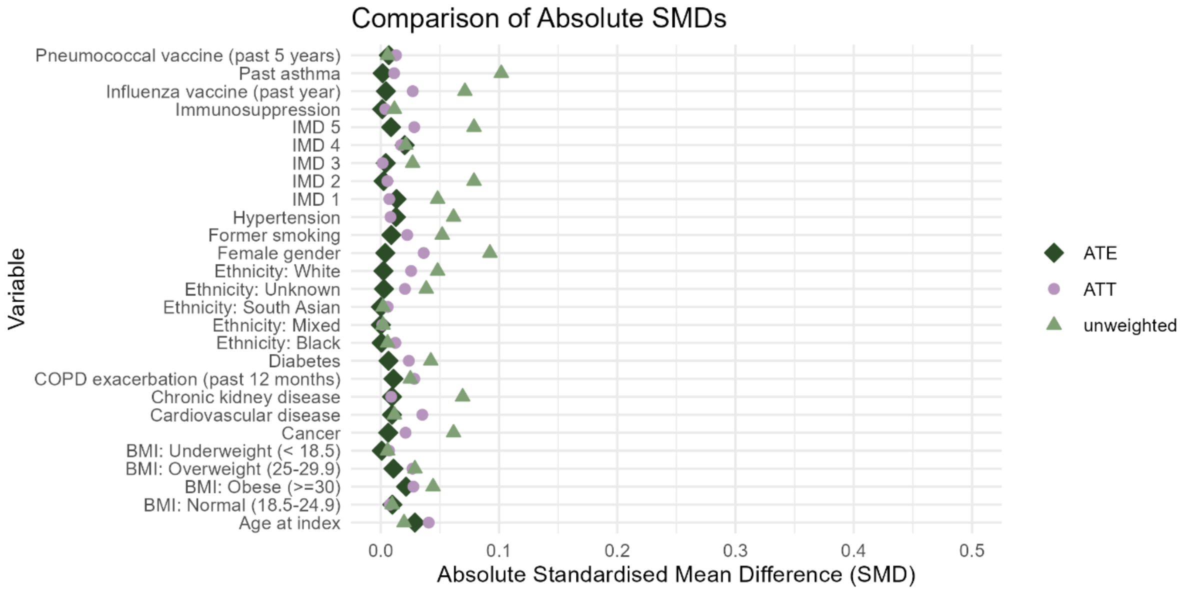

**Supplementary Method 1 Example calculation, scenario 1**

The odds of death in the hospitalised for each treatment group is as follows:

$$odds_{D=1|E=1,H=1} = \frac{n_{D=1,E=1,H=0}}{n_{D=0,E=1,H=0}} = \frac{42}{65} = 0.65 \quad (1)$$

$$odds_{D=1|E=0,H=1} = \frac{n_{D=1,E=0,H=0}}{n_{D=0,E=0,H=0}} = \frac{50}{83} = 0.60 \quad (2)$$

We assume the odds of death are the same among the non-hospitalised compared to the hospitalised (scenario 1).

As we have data on the number of COVID-19 deaths outside of hospitals by treatment group, we calculate the number of patients with severe COVID-19 who recovered outside of hospital.

$$n_{D=0,E=1,H=0} = \frac{n_{D=1,E=1,H=0}}{odds_{D=1,E=1,H=1}} = \frac{20}{0.65} = 30.95 \approx 31 \quad (3)$$

$$n_{D=0,E=0,H=0} = \frac{n_{D=1,E=0,H=0}}{odds_{D=1,E=0,H=1}} = \frac{22}{0.60} = 36.52 \approx 37 \quad (4)$$

Adding together the observed hospitalisations, the COVID-19 deaths without hospitalisation and the assumed number of recoveries without hospitalisation, we have 158 patients with severe COVID-19 in the ICS group and 192 in the LABA/LAMA group.

Having calculated the number of people with severe COVID-19 who recovered, we can calculate an odds ratio accounting for the people we did not observe in the hospitalisation data.

$$OR = \frac{n_{D=1,E=1} * n_{D=0,E=0}}{n_{D=1,E=0} * n_{D=0,E=1}} = \frac{62 * (83 + 37)}{72 * (65 + 31)} = 1.07 \quad (5)$$

Supplementary Table 1 2x2 table of hospitalisation and death for ICS group

| ICS group | | Hospitalisation | | |
|---|---|---|---|---|
| | | Hospital | No hospital | |
| Death | Death | $n_{D=1, H=1}$ = 42 | $n_{D=1, H=0}$ = 20 | $n_{D=1}$ = 62 |
| | Survived | $n_{D=0, H=1}$ = 65 | $n_{D=0, H=0}$ = ? | $n_{D=0}$ = ? |
| | | $n_{H=1}$ = 107 | $n_{H=0}$ = ? | |

Supplementary Table 2 2x2 table of hospitalisation and death for LABA/LAMA group

| LABA/LAMA group | | Hospitalisation | | |
|---|---|---|---|---|
| | | Hospital | No hospital | |
| Death | Death | $n_{D=1, H=1}$ = 50 | $n_{D=1, H=0}$ = 22 | $n_{D=1}$ = 72 |
| | Survived | $n_{D=0, H=1}$ = 83 | $n_{D=0, H=0}$ = ? | $n_{D=0}$ = ? |
| | | $n_{H=1}$ = 133 | $n_{H=0}$ = ? | |

Supplementary Table 3 Diagnostic checks of scenarios 1-4. For totals, decimals were rounded up to the nearest integer.

| | ICS | | | | | LABA/LAMA | | | | |
|---|---|---|---|---|---|---|---|---|---|---|
| Scenario | Odds of death among non-hospitalised | n (severe COVID-19, not hospitalised, recovered) | n (severe COVID-19, not hospitalised) | n (severe COVID-19) | p (hospitalisation) | Odds of death among non-hospitalised | n (severe COVID-19, not hospitalised, recovered) | n (severe COVID-19, not hospitalised) | n (severe COVID-19) | p (hospitalisation) |
| 1 | 0.65 | 31 | 51 | 158 | 0.68 | 0.60 | 37 | 59 | 192 | 0.69 |
| 2 | 1.29 | 16 | 36 | 143 | 0.75 | 0.60 | 37 | 59 | 192 | 0.69 |
| 3 | 0.32 | 62 | 82 | 189 | 0.57 | 0.60 | 37 | 59 | 192 | 0.69 |
| 4 | 1.29 | 16 | 36 | 143 | 0.75 | 1.20 | 19 | 41 | 174 | 0.77 |
